# Effects of maternal exercise on infant mesenchymal stem cell mitochondrial function, insulin action, and body composition in early infancy

**DOI:** 10.1101/2024.03.04.24303710

**Authors:** Filip Jevtovic, Donghai Zheng, Alex Claiborne, Ericka M. Biagioni, Breanna L. Wisseman, Polina M. Krassovskaia, David N. Collier, Christy Isler, James E. DeVente, P. Darrell Neufer, Joseph A. Houmard, Linda E. May

**Author notes:** Corresponding author **Contact Info:** Linda E May, Ph.D. East Carolina University Greenville, NC 27858.

## Abstract

**Objective:** Rates of pediatric obesity are continuously rising and are likely to translate into a high incidence of metabolic disease later in life. Maternal exercise (ME) has been established as a useful non-pharmacological intervention to improve infant metabolic health; however, mechanistic insight behind these adaptations remains mostly confined to animal models. Infant mesenchymal stem cells (MSCs) give rise to infant tissues (e.g., skeletal muscle), and remain involved in mature tissue maintenance. Importantly, these cells maintain metabolic characteristics of an offspring donor and provide a model for the investigation of mechanisms behind infant metabolic health improvements.

**Methods:** We used undifferentiated MSC to investigate if ME affects infant MSC mitochondrial function and insulin action, and if these adaptations are associated with lower infant adiposity.

**Results:** We found that infants from exercising mothers have improvements in MSC insulin signaling are related to higher MSC respiration and fat oxidation, and expression and activation of energy-sensing and redox-sensitive proteins. Further, we found that infants exposed to exercise in utero were seemingly leaner at 1-month of age, with a significant inverse correlation between infant MSC respiration and infant adiposity at 6-months of age.

**Conclusion:** These data suggest that infants from exercising mothers are relatively leaner and this is associated with higher infant MSC mitochondrial respiration, fat use, and insulin action.

## Introduction

The prevalence of pediatric obesity is escalating worldwide ^1^. The Developmental Origins of Health and Disease hypothesis suggests that the programming of metabolic disease begins *in utero* and is dependent on the environmental exposures experienced during intrauterine life. Observational human and animal studies have confirmed that suboptimal maternal nutrition promotes the transgenerational propagation of metabolic disease (i.e., obesity), partially through alterations of offspring substrate metabolism ^2–9^. Central to energy metabolism, and presumably involved in intergenerational health transmission, mitochondria display prominent plasticity and responsiveness to various physiological and pathophysiological stimuli.

Mesenchymal stem cells (MSCs) contribute to the fetal development of several tissues including skeletal muscle, and have been shown to align with the metabolic phenotype of the offspring donor ^2–5,10–14^. Further, these progenitor cells are evident in the developed tissue (i.e., satellite cells) and contribute to tissue maintenance. Maternal obesity has been linked to lower infant MSC fatty acid oxidation capacity, lower AMPK activity, and fatty acid partitioning towards complete rather than incomplete oxidation ^2,4^. These mitochondrial alterations are in parallel with increased infant adiposity and umbilical cord blood insulin levels and are contingent on maternal metabolic health. This showcases the susceptibility and responsiveness of mitochondrial biology to prenatal influences and could represent an early marker of underlying metabolic disease risk.

In non-gravid adults, aerobic and resistance exercise improves insulin action and mitochondrial function ^15–19^. While previous findings support the potential of maternal exercise (ME) to reduce infant adiposity and childhood obesity ^20^, the underlying biological mechanisms involved remain largely unexplored. Accordingly, we sought to elucidate if ME-induced improvements in infant adiposity are associated with the increase in infant MSC insulin action and mitochondrial capacity. Therefore, the primary purpose of this study was to determine if ME improves infant undifferentiated MSC insulin action, mitochondrial capacity, and fat metabolism. Additionally, we determined infant body composition measurements at 1- and 6-months of age, to examine whether changes in MSC mitochondrial functioning are associated with lower infant adiposity.

## Methods

### Ethics Statement

This study used umbilical cord MSCs and infant morphometrics collected in a randomized control trial (IRB#: 12-002524, ClinicalTrials.gov Identifier: NCT03838146). Approval for this study and all experiments was obtained from the East Carolina University Review Board and informed consent was obtained from each participant upon enrollment. Data from some of the participants in the current paper have been published previously^21^; however, the present paper includes data from participants collected since these previous publications, includes data from a 6-month infant follow up, and focuses on different aspects of MSC metabolism.

### Pre-intervention testing

Healthy females were recruited between 13-16 weeks of gestation based on the inclusion criteria outlined in the study design Figure 1. After receiving clearance from their obstetric provider, participants were randomly assigned (via sealed, sequentially numbered envelopes derived from computer-generated randomization, Graph-Pad Prism Software) to aerobic, resistance, combination (aerobic+resistance) exercise, or non-exercising control group.

**Figure 1.**
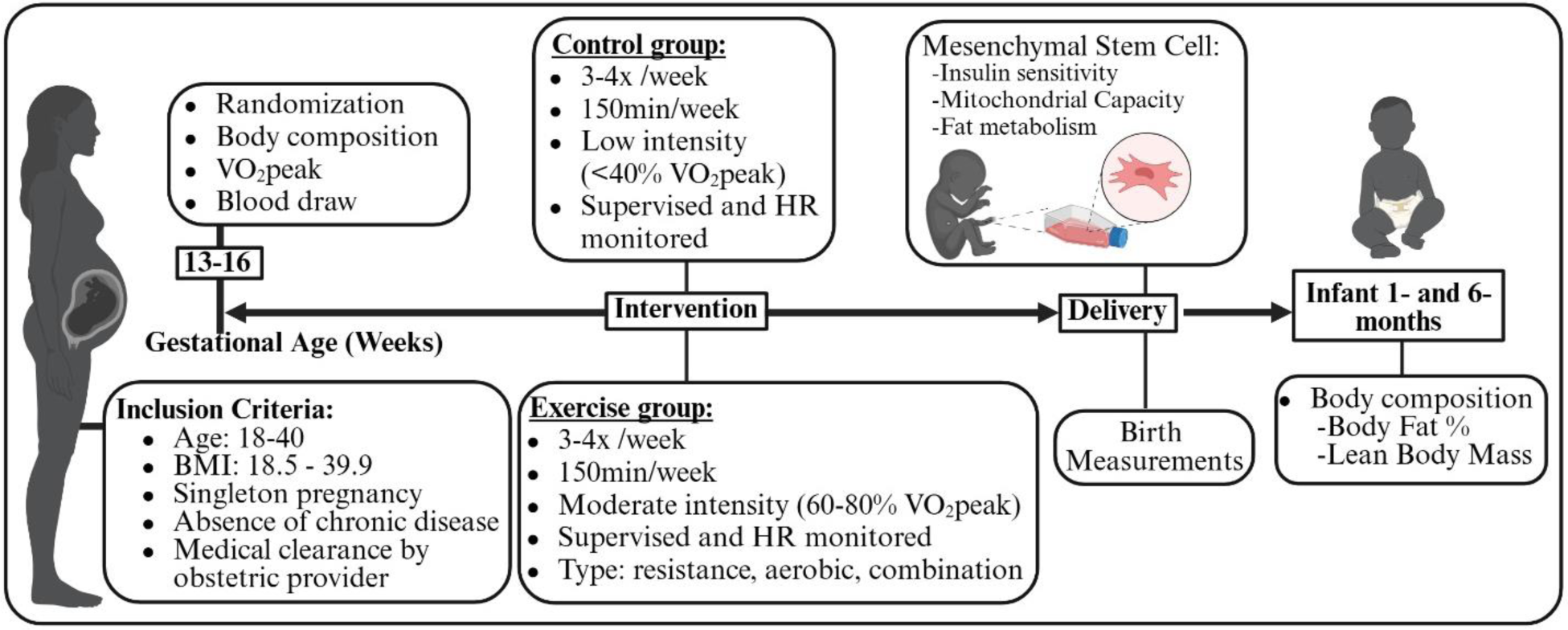
Design and timeline of the study. Created with BioRender.

Participants recruited before or after COVID-19 pandemic completed a submaximal modified Balke treadmill test following previously described submaximal modified Balke treadmill test ^22^ to measure peak oxygen consumption (VO_2peak_), and to determine target heart rate (THR) zones for subsequent exercise training. THR zones corresponded to maternal HR at 60-80% of maximal oxygen consumption, reflecting moderate intensity ^22^. For participants recruited during COVID-19 pandemic, to minimize exposure and potential risk associated with exercise testing, THR were determined based on the pre-pregnancy physical activity level and age, using published guidelines^22^.

### Exercise Intervention

As published previously ^23^, participants exercised according to American College of Obstetricians and Gynecologists guidelines for the duration of their pregnancy (∼16-40 weeks). Participants performed moderate intensity (60%-80% maximal oxygen consumption and 12-14 rated perceived exertion) aerobic, resistance or a combination exercise. Every session was supervised. Additionally, we used HR monitors to ensure that participants exercised within their THR zone, corresponding to moderate intensity, regardless of the training mode. The control group performed supervised stretching, breathing, and flexibility exercises at low intensity (<40% VO2peak). ME adherence was calculated by dividing the number of sessions attended by the total number of possible sessions within the participants’ gestational period. ME intensity (METs) was based on the published compendium of physical activity for the exercise performed each session^24^. Average ME dose during each week, expressed as MET·min/wk, was quantified (frequency X duration of session) then multiplied by the intensity (METs) of their exercise. Further, total volume of exercise during pregnancy (Total MET·min) was calculated by multiplying the MET·min/wk by the total number of weeks of gestational exercise. Importantly, average, and total exercise volumes were calculated for 20 weeks between 16-36 weeks of gestation, to avoid the influence of different gestational length between mothers (38-41 weeks) on exercise volume. The ability of our protocol in eliciting maternal physiological adaptations to exercise has been previously reported ^25^.

### Maternal Measurements

Maternal measurements were obtained as previously described ^23^. Maternal age, parity, pre-pregnancy weight and height and body mass index (BMI, kg/m^2^), gestational diabetes mellitus status (yes or no), 1-hour glucose value during an oral glucose tolerance test (OGTT), gestational weight gain (GWG), length of gestation, mode of delivery, and breastfeeding status, were abstracted from various sources including pre-screening eligibility and postpartum questionnaires as well as maternal and neonatal electronic health records. At 16 weeks of gestation, we obtained maternal BMI and determined maternal percent body fat using a previously validated skinfold technique and age-adjusted equations ^26,27^. Additionally, maternal fingerstick blood was analyzed using Cholestech LDX Analyzer (Alere Inc., Waltham, MA, USA) and point of care Lactate Plus Analyzer (Nova Biomedical, Waltham, MA, USA) for the quantification of maternal lipids (total cholesterol (TC), triglycerides (TG), HDL, non-HDL, LDL), glucose and lactate.

### Infant Measurements

Birth measurements (weight, length, BMI, abdominal, head and chest circumference, 1 and 5-minute Apgar scores) and infant sex were extracted from neonatal electronic health records. At one and six months of age, infant weight (kg), length (m), BMI (kg/m^2^), and body fat (%), were measured by trained staff in a designated pediatric clinic. Weight and length were measured using a standard, calibrated infant scale and horizontal stadiometer, respectively. Skinfold thickness was measured via calibrated Lange calipers at three designated anatomical sites on the right side of the infant’s body: biceps, triceps, and subscapular. Values for these sites were summed to determine skinfold thickness. The skinfold thickness data was then used to calculate the percent body fat (BF) using the following equation by Slaughter et al.^28^:

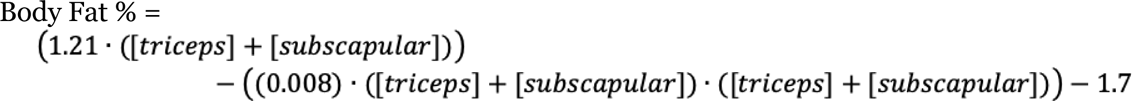

Fat-free mass was calculated using the following two-step equation ^2^^9^:

1st step:

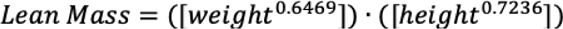

2nd step:

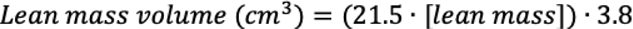

Due to participant attrition, data was available for 33 (Control=6, Exercise=27) infants at 1-month and 17 (Control=3, Exercise=14) infants at a 6 month follow up.

### MSC Methods

*Isolation*. MSC isolation followed previously published protocols ^13,14,21,30^. Briefly, a 4-inch section was cleared of blood and vessels, and dissected into ∼0.5-cm2 pieces and placed Wharton’s jelly/lumen side-down onto droplets of 30% BSA in 10-cm2 dishes. The tissue sections were incubated in mesenchymal growth media containing low glucose DMEM (Gibco laboratories, Grand Island, NY) supplemented with 10% MSC-qualified fetal bovine serum (FBS), 1x Gentamicin Amphotericin (Life Technologies, Gaithersburg, MD), until cells reached 80% to 100% confluency, at which point they were cryopreserved in liquid nitrogen. All experiments were completed in undifferentiated MSCs in passages 3-5.

### Fatty Acid Metabolism

For the assessment of natural lipid content cells were rinsed with DPBS, fixed with 4% paraformaldehyde, and stained using 0.5% Oil Red O stain (ORO) in propylene glycol. Thereafter, cells were rinsed with 85% propylene glycol and deionized water, and ORO stain was solubilized with isopropanol. Degree of ORO staining was analyzed spectrophotometrically at 520 nm. To account for cell density, each well was then stained with 0.3% Janus Green. After rinsing with deionized water, cellular Janus Green was solubilized with 0.5 M HCl; samples were analyzed spectrophotometrically at 595 nm for degree of Janus Green staining. Cellular Oil Red O stain was then normalized to cellular Janus Green stain. All staining was done with 5 technical replicates.

For fatty acid oxidation, following 3 hours of serum starvation MSCs were washed with DPBS and incubated in ^14^C-oleate-containing media (d-[1-^14^C] oleate (Perkin-Elmer; 1.0 µCi/mL, 5.0 mM glucose, 200 µM oleate, 1 mM carnitine) for 2 hours at 37 °C. The experimental media were transferred into a customized 48-well trapping plate with fabricated grooves between 2 continuous wells. CO_2_ in the media was acid trapped in 1N NaOH via the addition of 70% perchloric acid. Oxidation of ^14^C-oleate into CO_2_ was determined with liquid scintillation counting of the conditioned 1N NaOH to derive a rate of complete fatty acid oxidation. Acidified media was collected following CO_2_ trapping and centrifuged at 12,000 x *g* at 4°C to determine levels of ^14^C-acid soluble metabolites (ASM) (i.e., acyl-carnitine, acetyl-carnitine, acetyl-coenzyme A, and short-chain fatty acyl-coenzyme A) as described previously ^13,14,21,30^. Data were corrected for total protein content and quantified via bicinchoninic acid (BCA) assay (Pierce Biotechnology, Inc.).

### Glucose Oxidation and Glycogen Synthesis

Glucose oxidation and glycogen synthesis were measured as described previously ^13,14,21,30^. In brief, following 3 hours of serum starvation, MSCs were incubated with media containing d-[1-^14^C] glucose (Perkin-Elmer, MA, USA; 1.5 µCi /mL, 5.0 mM glucose) in the presence (100nM) or absence of insulin at 37 °C for 2 hours. Complete glucose oxidation to CO_2_ was determined following the same protocol as described above for fatty acid oxidation. Rate of nonoxidized glycolytic metabolite production (e.g., lactate, pyruvate), indicative of the rate of non-oxidized glycolysis (NOG), was measured by putting unacidified media collected during the glucose oxidation experiments onto ion-exchange cellulose paper (anion exchange paper; Macherey-Nagel, Duren, Germany), followed by 30 minutes of drying and 4 washes of 10 minutes each with dH2O. After washing, 14C-labeled glucose incorporation into nonoxidized glycolytic metabolites was determined via liquid scintillation counting of the cellulose paper. Finally, MSCs were then washed twice with DPBS and solubilized with 0.5% SDS. Part of the lysate was combined with carrier glycogen (1 mg), denatured with heat, and precipitated overnight using 100% ethanol. Once precipitated, glycogen pellets were washed with 70% ethanol to eliminate residual glycogen-unbound glucose or glucose derivatives (i.e., G-6-P), resuspended with dH_2_O, and the incorporation of radioactive glucose into glycogen was determined with liquid scintillation counting. All data was normalized to protein using BCA assay.

### Immunoblotting and citrate synthase activity

MSCs were incubated in the presence of absence of 100nM of insulin for 10 minutes. MSCs were rinsed with DPBS and lysed in ice-cold mammalian protein extraction reagent (M-PER; Thermo Scientific Waltham, MA) containing phosphatase (2 and 3) and protease inhibitor cocktails (Sigma, St. Louis, MO), 10 mM sodium orthovanadate, and 1% Triton (Bio-Rad, Hercules, CA). Samples were sonicated for 5 s, then rotated end-over-end at 4 °C for 2 h and centrifuged at 12,000 × g at 4 °C for 15 min. The supernatant was collected, and protein concentrations were determined (BCA Assay). Cell lysates were mixed with 4x Laemmli Sample Buffer (Bio-Rad, Hercules, CA), separated by SDS-PAGE electrophoresis, and transferred to the nitrocellulose membrane using Trans-Blot® TurboTM Transfer System (Bio-Rad). Membranes were blocked with 5% bovine serum albumin (BSA) in TBS-T (1x TBS, 0.1% Tween-20) for 1 h, then incubated overnight at 4 °C with primary antibodies (see Supplementary Table 1) diluted in 5% BSA in TBS-T. Secondary antibodies (LI-COR, Cat. #s 926-32211, 926-32210, 926-68072) were diluted in 5% BSA in TBS-T (1x TBS, 0.1% Tween-20). All data were normalized to β-Actin protein expression. Citrate synthase activity was determined using colorimetric plate-based assay and previously published methods^31^, and normalized to content.

### Cellular Respirometry

High-resolution O_2_ consumption measurements were conducted using the Oroboros Oxygraph-2K (Oroboros Instruments, Innsbruck, Austria) (O2K) in intact and digitonin permeabilized cells, using intact cell respiration media (1xDMEM power without HCO_3_, 20mM HEPES, and MSC FBS) or O2K assay buffer (Potassium-MES (105mM; pH = 7.1), KCl (30mM), KH_2_PO_4_ (10mM), MgCl_2_ (5mM), EGTA (1mM), BSA (0.5mg/ml), 5mM creatine monohydrate) respectively. <u>Intact cell</u> <u>respiration:</u> after recording the basal respiration, maximal respiratory capacity of intact cells was assessed by addition of the uncoupler FCCP (Carbonyl cyanide p-trifluoro-methoxyphenyl hydrazone, C_10_H_5_F_3_N_4_O) titration (0.5-3.5µM). <u>Permeabilized cell assays:</u> cells were permeabilized in chamber with digitonin (0.01 mg/mL), and saturating carbon substrates (pyruvate (P) 5mM, malate (M) 1mM, glutamate (G) 5mM, succinate (S) 5mM, octanoyl-l-carnitine (O) 0.2mM) were added. Maximal coupled and uncoupled respiration was assessed by the addition of ADP (0.5mM) and FCCP (0.5-3.5µM), respectively. <u>Complex I, II, and IV maximal respiration:</u> was assessed in permeabilized cells under saturating ADP and sequential additions of P/M (complex I); rotenone (2mM, Complex I inhibitor) and S (10mM) for complex II; and malonate (20mM, complex II inhibitor), antimycin A (2µM, complex III inhibitor), and TMPD (N,N,N’,N’-tetramethyl-p-phenylenediamine) and ascorbate (0.5mM each) for complex IV. C<u>reatine kinase (CK) clamp</u> ^31^ was used to determine steady-state oxygen consumption across a range of cellular energetic states ΔG_ATP_, -12.94 kcal·mol^-1^ (high energetic demand) and -14.7 kcal·mol^-1^ (low energetic demand), using known amounts of creatine (Cr), phosphocreatine (PCr), and ATP in the presence of excess amounts of CK. After permeabilization, CK clamp was established by the addition of ATP (5 mM), PCr (1 mM), and CK (20U/ml), simulating a ‘maximal’ demand for ATP re-synthesis. Thereafter, sequential additions of P/M (5mM/1mM), cytochrome C (10 µM), and G/S/O (5 mM/5mM/0.2mM), were performed. Further, PCr additions (6, 9, 15, and 21mM) were performed to gradually return ATP demand toward baseline. Cytochrome C (10 µM) was used to check the integrity of the outer mitochondrial membrane in each experiment, and no increase in respiration relative to the pre-cytochrome C rate, was used as a quality control assessment for outer membrane integrity. Non-mitochondrial respiration was controlled by adding Ant A (2µM). Data were normalized to viable cell count and expressed as pmol/s/million cells.

### Statistical Analysis

Comparison between exercise groups (aerobic, resistance or combination) were performed using one way ANOVA, ANCOVA, or two-way repeated measure ANCOVA. Considering that MSC phenotype was similar across exercise groups (Supplementary Table 2, Figures 1-5), exercise groups were combined in one ‘ME’ group to increase sensitivity to detect the effects of ME on different measurements. Thereafter, comparison between control and combined exercise group was performed using unpaired parametric and nonparametric t-tests, ANCOVA, and two-way repeated measures ANCOVA, where appropriate. Factors tested were group (control vs exercise) and insulin (basal vs insulin). Bonferroni post-hoc test was performed when significance was detected in either main effects or interactions. Pearson’s correlations were used to determine if there were any relationships between variables. Statistical significance was set at *p* ≤ 0.05. Statistical analyses were performed using GraphPad Prism version 9.3 (GraphPad Software, San Diego, CA) for Windows.

## Results

Twelve of the 41 women started the trial during the COVID-19 pandemic and had their THR zones estimated based on the previously established protocols ^22^; however, MSC outcomes were similar between subjects that were assigned exercise intensity based on actual vs. estimated HR (p>0.05).

Average adherence for exercisers was 90±7% (mean ± SD). Maternal and infant characteristics are presented in Table 1. Compared to control subjects, exercisers had higher VO_2_ peak (p<0.01) and have exercised significantly more during pregnancy (MET*min/wk, p<0.001; Total MET*min, p<0.001). Further, exercisers had lower self-reported pre-pregnancy BMI (p≤0.05) than controls; however, BMI and body fat (%) were similar between groups at 16-weeks of gestation. Maternal lipids and lactate were similar across groups, and all mothers had normal fasting glucose <99 mg/dL. Further, all OGTT measurements were within normal range (<140 mg/dL) indicating similar degree of glucose tolerance across groups and absence of gestational diabetes. Across groups, infants had similar sex distribution, birth weight, height, BMI, Apgar scores, and all were breastfed. Considering the difference in pre-pregnancy BMI between groups, this measure was included as a covariate in subsequent analysis. We did not include blood glucose as a covariate because all participants had normal fasting and 1 hour glucose during OGTT, and blood glucose was not significantly correlated with any of the MSC or infant outcomes.

**Table 1.** Maternal and Infant Characteristics.

| **Table 1. Maternal and Infant Characteristics** |  |  |  |
| --- | --- | --- | --- |
| **Maternal Characteristics** | **Control (n=11)** | **Exercise (n=30)** | **p-value** |
| Age (yrs) | 28.3±5.3 | 30.4±3.5 | 0.15 |
| VO<sub>2</sub>peak (ml/kg/min)* | 18.3±3.9 | 24.4±4.8 | **0.01** |
| Average MET*min/wk between 16-36 weeks of gestation | 134±134 | 573±97 | **<0.001** |
| Total MET*min between 16-36 weeks of gestation | 1881±2071 | 13068±2782 | **<0.001** |
| Pre-pregnancy BMI | 28.3±4.8 | 24.6±3.9 | **0.02** |
| 16-week BMI | 28.9±5.0 | 25.9±4.3 | 0.06 |
| 16-week Body Fat (%) | 34.2±5.8 | 34.0±4.1 | 0.89 |
| 16-week Total Cholesterol (mg/dl) | 179.4±32.7 | 176.9±31.2 | 0.83 |
| 16-week LDL (mg/dl) | 92.6±28.8 | 94.8±29.5 | 0.85 |
| 16-week HDL (mg/dl) | 65.9±14.0 | 62.2±16.1 | 0.56 |
| 16-week non-HDL (mg/dl) | 109.3±26.7 | 114.9±31.6 | 0.65 |
| 16-week Triglycerides (mg/dl) | 88.8±41.1 | 100.1±36.6 | 0.42 |
| 16-week Glucose (mg/dl) | 84.2±7.4 | 78.6±6.4 | **0.03** |
| 16-week Lactate (mmol/l) | 1.1±0.6 | 0.9±0.5 | 0.46 |
| OGTT (mg/dL) | 113.0±21.7 | 108.0±27.3 | 0.64 |
| Gestational weight gain (kg) | 14.0±4.8 | 16.5±5.6 | 0.20 |
| Gestational length (weeks) | 38.9±0.9 | 39.6±1.2 | 0.09 |
| Parity | 1 (0, 2) | 0 (0, 3) | 0.07 |
| Mode of delivery (SVD/C-section) | 6/5 | 22/8 | 0.96 |
| **Infant Characteristics** | **Control (n=11)** | **Exercise (n=30)** | **p-value** |
| Fetal sex (F/M) | 3/8 | 9/21 | 0.99 |
| Birth weight (kg) | 3.7±0.5 | 3.5±0.4 | 0.16 |
| Birth length (m) | 0.50±0.02 | 0.49±0.02 | 0.61 |
| Birth BMI | 15.0±2.0 | 14.3±1.7 | 0.26 |
| Head circumference (m) | 0.35±0.02 | 0.34±0.02 | 0.35 |
| Chest circumference (m) | 0.34±0.2 | 0.33±0.02 | 0.60 |
| Abdominal circumference (m) | 0.32±0.02 | 0.31±0.02 | 0.48 |
| Apgar-1 minute | 8 (2, 9) | 8 (7, 9) | 0.99 |
| Apgar-5 minute | 9 (8, 9) | 9 (9, 9) | 0.53 |
| All data expressed as mean ± SD, t-test, p≤0.05. * VO<sub>2</sub>peak n=29/41 due to COVID; OGTT = 1-hour blood oral glucose tolerance test at 24-28 weeks; Gestational weight gain = delivery - pre-pregnancy; SVD = spontaneous vaginal delivery; F= female infant sex; M = male infant sex. |  |  |  |

### ME improves infant MSC insulin action

Phosphorylation of Akt was similar between groups in the non-insulin stimulated (basal) state (Figure 2A-B). Both groups increased Akt activation upon insulin exposure (p<0.0001); however, MSCs of exercise-exposed (Ex-MSC) compared to non-exercise-exposed infants (Ctrl-MSC) had a higher phosphorylation of Akt at both Ser473 and Thr308 (p≤0.05). Additionally, there was a treatment by group interaction for Akt phosphorylation at Ser473 (p≤0.05), but only trending interaction for Thr308 (p=0.1). Further, while absolute rates of glycogen synthesis in the basal and insulin-stimulated conditions were similar across groups, there was a greater relative increase in glycogen synthesis rate upon insulin exposure in Ex-MSCs (32%) compared to Ctrl-MSCs (20%) (group*treatment, p≤0.05, Figure 2C). Finally, Ex-MSCs had significantly higher relative phosphorylation of AMPK (p≤0.05), and PGC-1α expression (p≤0.05), as well as a trend towards higher SIRT1 content (p=0.073) compared to CTRL-MSCs (Figure 2D). Together, these data indicate improvements in MSC insulin action with ME which is associated with the upregulation of energy and redox-sensitive proteins.

**Figure 2.**
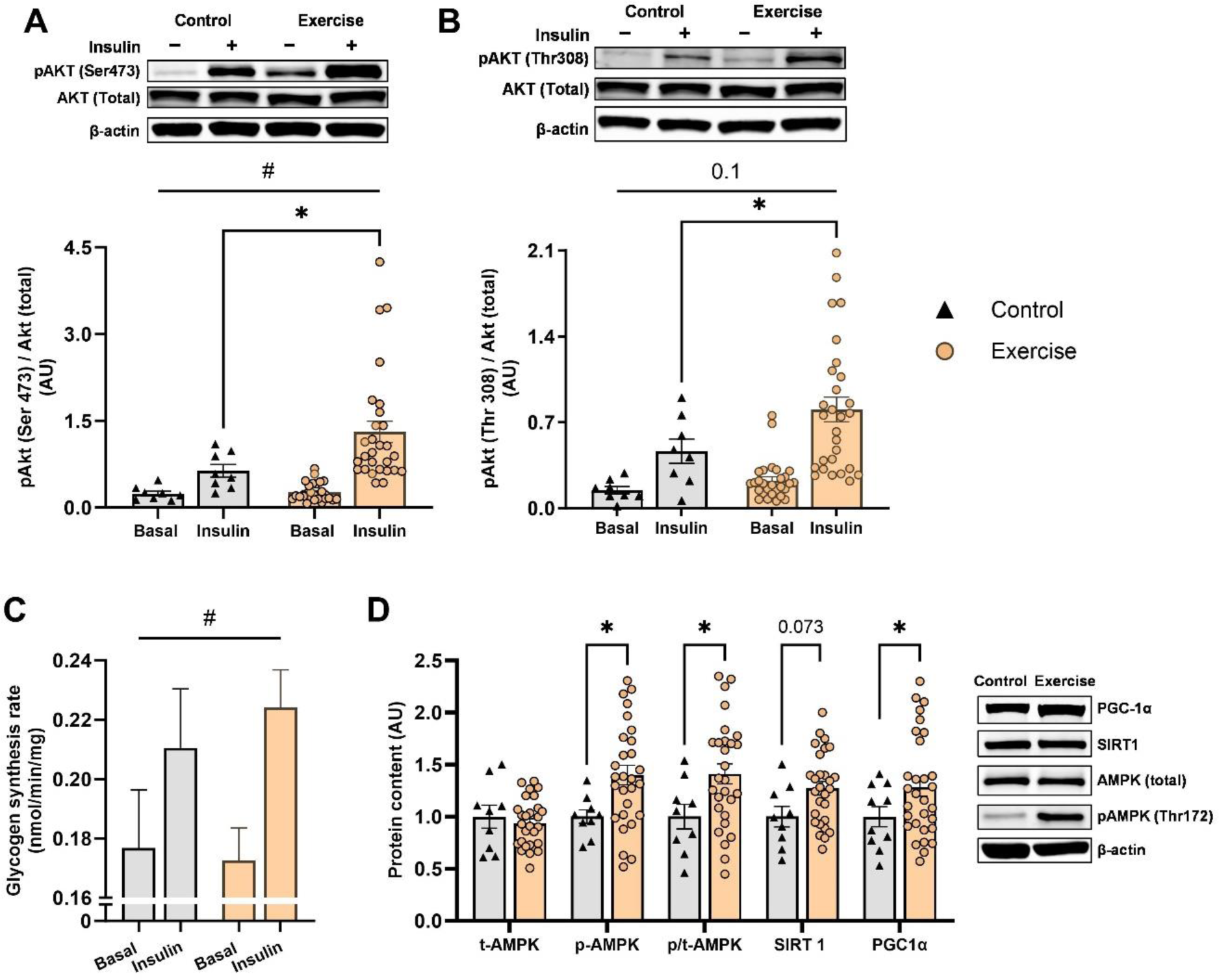
Ex-MSC display higher insulin action compared to Ctrl-MSCs. Western blot analysis shows the ME-induced increases in insulin-stimulated phosphorylation of Akt at Ser473 (A) and Thr308 (B) residues. Higher response to insulin stimulation was observed for Ser473 activation (A) and glycogen synthesis (C) in Ex-MSCs compared to Ctrl-MSCs (group*treatment interaction p≤0.05). Energy and redox-sensitive protein expression and activation was higher in Ex-MSCs (D). Insulin signaling and glycogen synthesis data were assessed by two-way repeated measures ANCOVA with Bonferroni post hoc test for the main effect (Ex-MSC vs Ctrl-MSCs). Ctrl-MSCs (n=8-11), Ex-MSCs (n=28-30). Mean±SEM *p≤0.05 for difference between groups. # p≤0.05 group*treatment interaction.

### ME increases intact MSC respiration

Ex-MSC compared to Ctrl-MSCs had higher intact cell basal and maximal respiration (p≤0.01, Figure 3A). Both groups had a similar increase in respiration from basal to maximal state (p>0.05 Figure 3B). We found similar citrate synthase activity (p>0.05, Figure 3C), a commonly used mitochondrial content surrogate, as well as a similar OXPHOS complex and pyruvate dehydrogenase protein expression between groups (p>0.05, Figure 3D-E). Interestingly, citrate synthase protein expression was greater in exercise compared to control MSCs (p≤0.05, Figure 3E). Together, Ex-MSCs display an elevated intact cell respiration that is seemingly independent of any differences in cell mitochondrial content.

**Figure 3.**
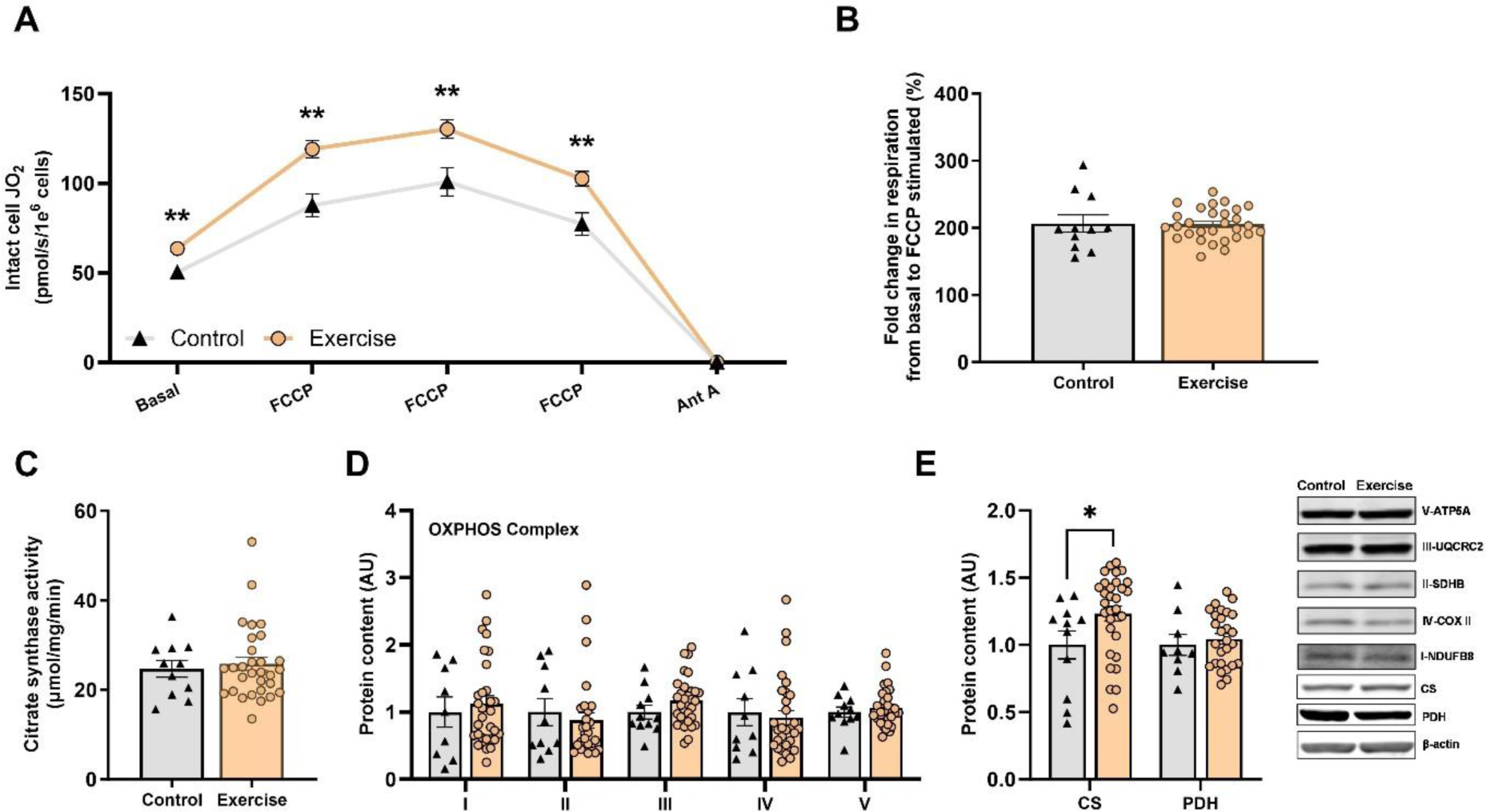
ME increases infant MSC respiration but not OXPHOS content. Ex-MSCs display higher intact cell respiration in basal state and across a range of mitochondrial uncoupler titrations (A) compared to Ctrl-MSCs, yet have similar increase from basal to maximal respiration (B). Mitochondrial maximal citrate synthase activity (C) was similar between groups. Western blots were used to quantify the expression of mitochondrial oxidative phosphorylation (OXPHOS) proteins (D), citrate synthase (CS) and pyruvate dehydrogenase (PDH) (E). Ctrl-MSCs (n=9-11), Ex-MSCs (n=29-30); * p≤0.05 ** p≤0.01

### An increase in MSC respiration is independent of OXPHOS adaptations

To investigate if higher respiration in Ex-MSCs is a result of intrinsic OXPHOS remodeling, two complementary assays in digitonin permeabilized cells were performed. Use of permeabilized cells allows for direct access to mitochondria and eliminates cytosolic and membrane transport input. Assays were performed with saturating carbon substrates (P/M/G/S/O) so that maximal electron transport system flux can be achieved. Respiratory capacity (State 3 or 3u) was similar across groups (p>0.05, Figure 4A). A higher intact cell but similar mitochondrial respiratory capacity suggests a difference in regulatory control within the cytosol (or membrane transport) of MSCs between the two groups. A limitation of this assay is the lack of control over cellular adenylate pool, and thus energetic state (i.e., ΔG_ATP_).

**Figure 4.**
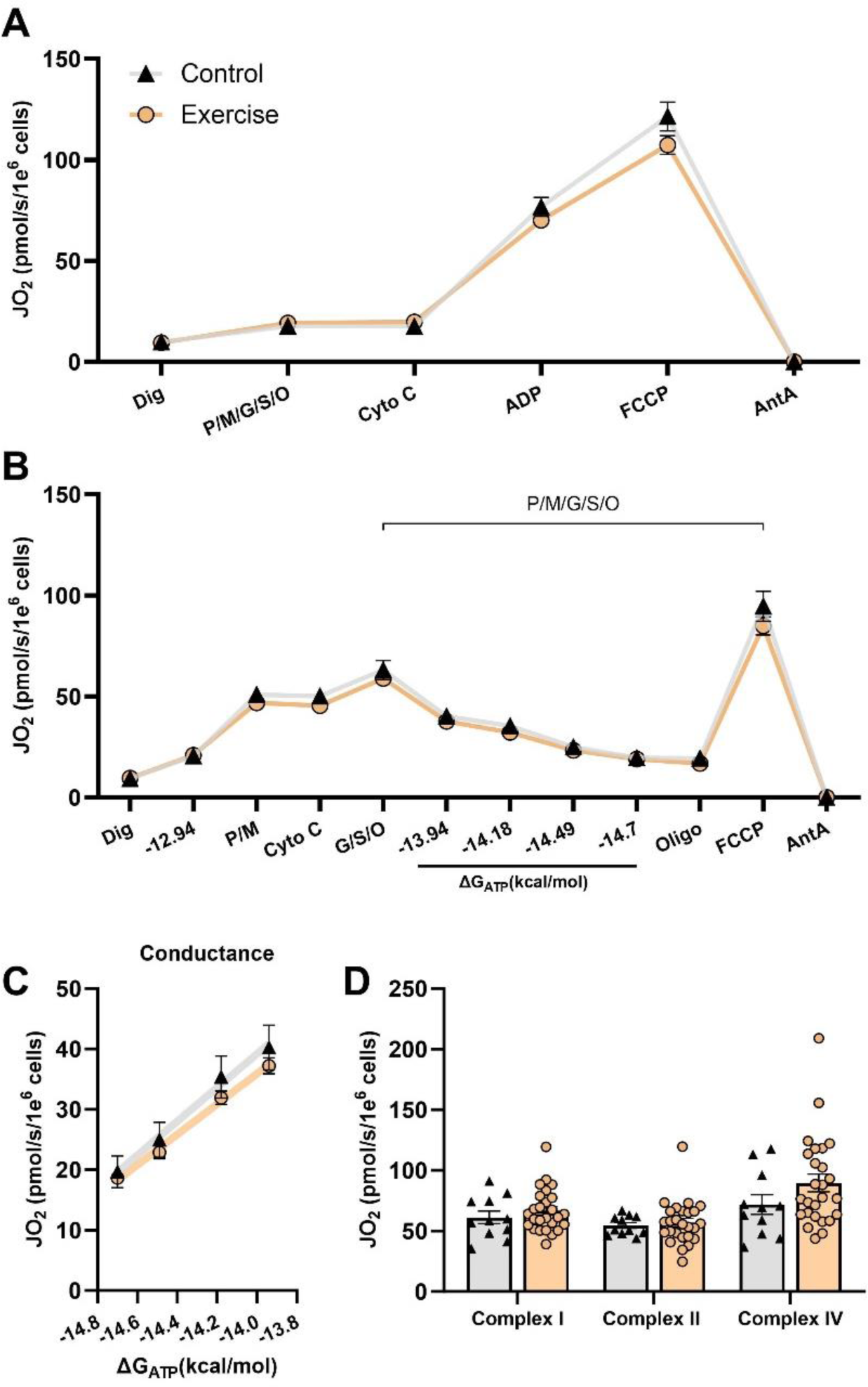
Permeabilized cell respiration is similar between groups. Permeabilized cell assays were used to gain direct access to mitochondria, void of cellular membrane transport and cytosol. Maximal oxygen consumption (JO_2_) in permeabilized cells (A), regardless of the presence of ΔG_ATP_ (B) was similar between groups. OXPHOS conductance (C). Maximal respiration supported with complex I, II, and IV specific substrates (D). Ctrl-MSCs (n=11), Ex-MSCs (n=29-30)

To address this, the creatine kinase clamp technique (Figure 4B) was employed to assess mitochondrial respiratory function over the entire range of ATP free energy (ΔG_ATP_; -12.9 (high) to -15.3 (low) kcal/mol). In this assay, the slope of the linear force-flow relationship between calculated ΔG_ATP_ and the corresponding steady-state oxygen consumption rate represents conductance, or sensitivity of the entire respiratory system to the changes in energy demand. There were no differences between groups in mitochondrial respiration or conductance (p>0.05, Figure 4B; p>0.05, Figure 4C). Finally, maximal respiration supported with complex I, II, and IV specific substrates was similar between groups (p>0.05, Figure 4D). Together, these data indicate that higher mitochondrial capacity in intact Ex-MSCs is independent of intrinsic mitochondrial OXPHOS adaptations.

### ME increases infant MSC lipid content and oxidation

Previous work has associated higher infant adiposity with lower MSC fat oxidation. Accordingly, we sought to test if ME-induced increases in infant MSC respiration are in parallel with changes in fatty acid oxidation and/or storage. Compared to Ctrl-MSCs, Ex-MSCs had ∼100% higher neutral lipid content (p<0.0001, Figure 5A). There were no differences between groups in mitochondrial lipid availability (p>0.05, Figure 5B), represented by a sum of complete (CO_2_) and incomplete fat oxidation (ASM), and indicative of all fatty acids taken up into the mitochondria during the measurement period. Of those lipids, Ex-MSCs had higher complete fatty acid oxidation to CO_2_ (p≤0.05), but similar incomplete fat oxidation rate (ASM production) (p>0.05, Figure 5B). Together, this difference between groups resulted in a higher partitioning ratio of incomplete (ASM) to complete (CO_2_) fatty acid oxidation in the Ex-MSC (p≤0.05, Figure 5B).

**Figure 5.**
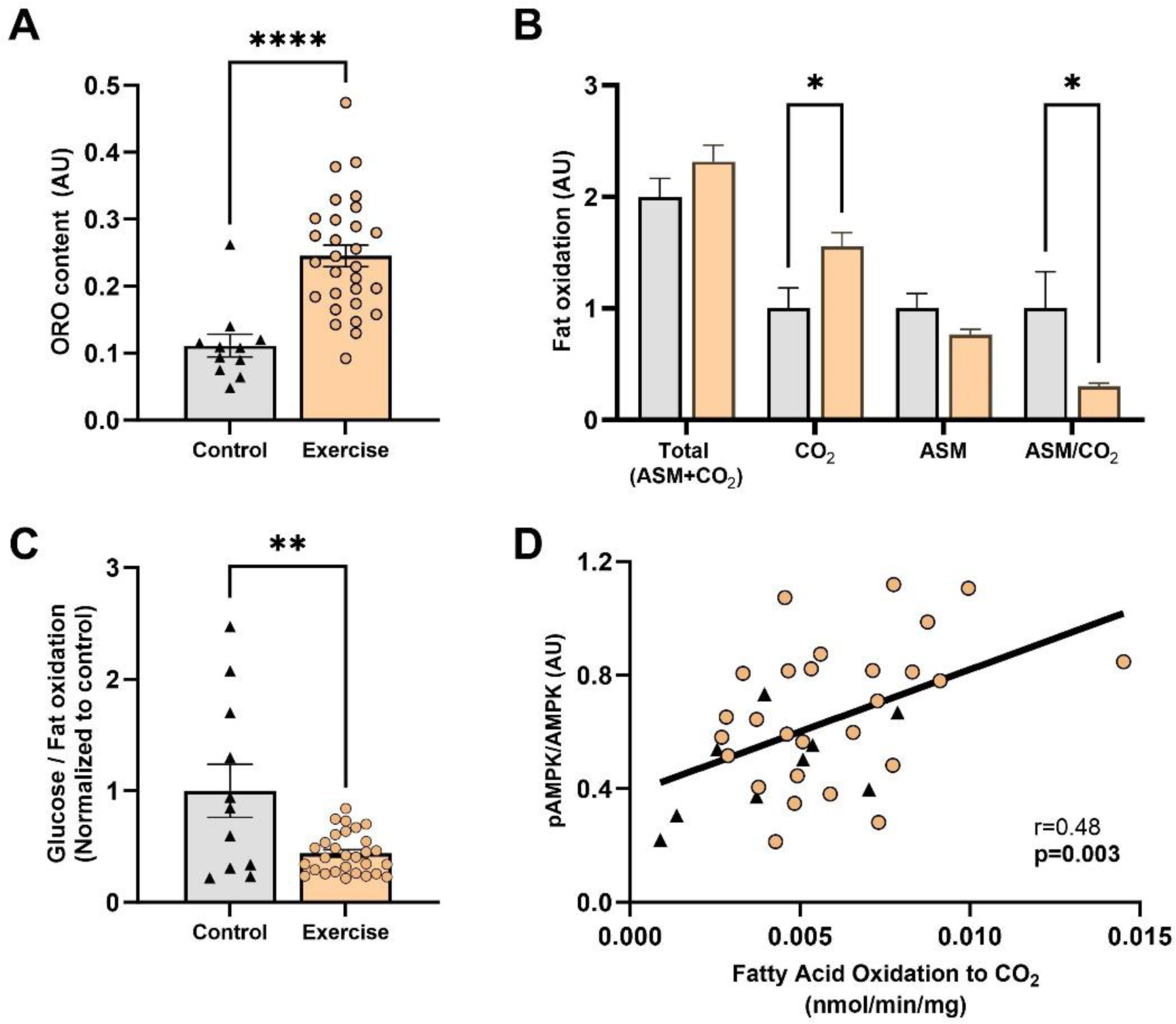
Ex-MSCs higher fat oxidation is associated with higher AMPK activation. ME increases neutral lipid content measured by Oil Red O staining (ORO) (A). Ex-MSCs have higher complete (CO_2_) and similar incomplete (ASM) fat oxidation (B). Higher ratio of glucose:fat oxidation in Ctrl-MSCs is indicative of lower fatty acid flux through TCA cycle (C). Correlation between AMPK activation and complete fat oxidation (D). All data expressed as mean±SEM; Ctrl-MSCs (n=11), Ex-MSCs (n=29-30). * p≤0.05 ** p≤0.01 **** p≤0.0001

To assess if there is a differential substrate oxidation, we additionally measured the rate of glucose oxidation. There was a similar glucose oxidation to CO_2_, and similar rate of non-oxidized glycolysis between groups (p>0.05, Supplementary figure 6). Interestingly, Ctrl-MSCs displayed a greater ratio of glucose to fat oxidation compared to Ex-MSCs (p<0.01, Figure 5C). Considering that fatty acid oxidation is measured in the presence of glucose, we interpret a lower glucose to fatty acid ratio in the Ex-MSC group as an indicator of an overall greater flux of fatty acid-derived acetyl-CoA through the TCA cycle in the Ex-MSCs. Finally, there was a significant direct correlation between complete fatty acid oxidation and AMPK phosphorylation (r=0.48, p<0.01, Figure 5D). Together, these data indicate that Ex-MSCs store and completely oxidized fat more than Ctrl-MSCs.

### Infants from exercisers are leaner at 1-month

At one month of age, infants from exercisers had significantly lower BMI and higher lean mass z-scores compared to infants from controls (p≤0.05, Figure 6A). Further, there was a trend towards lower infant body fat percentage in infants from exercisers at 1-one-month (p=0.1). Since infants experience a steep rate of adipose tissue expansion within the first 6 months of life ^32^, we did a six-month follow up with infants from both groups. Due to follow-up attrition rates, infant body fat percentage data was available for 17 infants (n=3 Control, n=14 Exercise), which prohibited group comparison. However, there was a significant inverse correlation between MSC basal (p<0.01, r=-0.62) and maximal respiration (p=0.01, r=-0.59) with six-month infant body fat percentage (Figure 6D-E). Together, these data show that ME decreases infant adiposity, and that lower adiposity is potentially linked to increases in infant MSC mitochondrial respiration.

**Figure 6.**
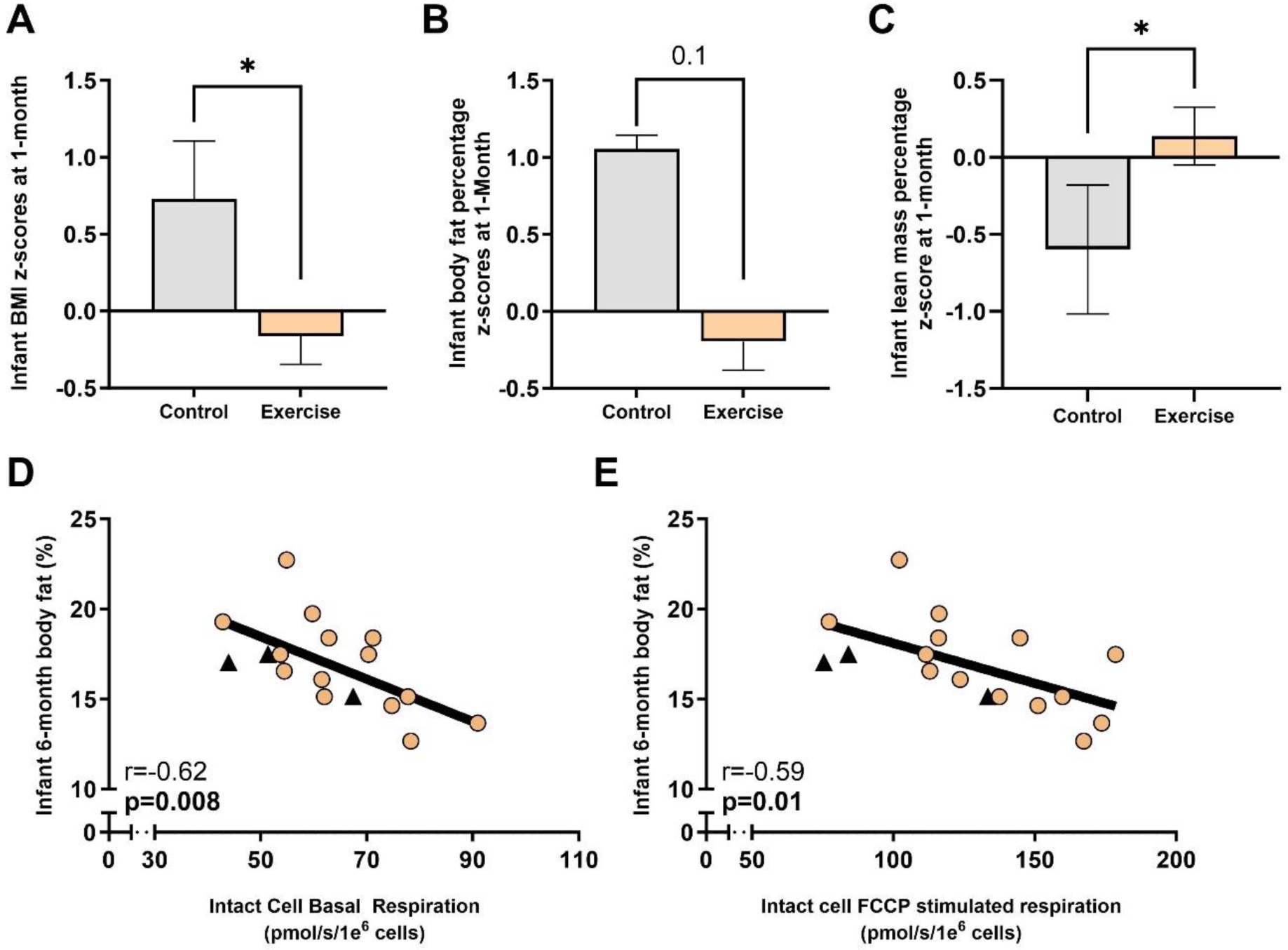
Infants from exercisers are seemingly leaner. At 1-month of age, infants from exercise group had significantly lower BMI z-scores (A) and a trend towards lower infant adiposity (B). Infant lean mass was significantly higher in exercise compared to control group (C). Higher intact cell basal and maximal (FCCP stimulated) respiration (JO_2_) was associated with lower infant adiposity at 6-month follow-up (D-E). All data expressed as mean±SEM; Ctrl-MSCs (n=6), Ex-MSCs (n=27) for 1 month comparison. Triangles -Ctrl-MSCs; Circles – Ex-MSCs. * p≤0.05

### ME volume during pregnancy is associated with infant MSC phenotype and adiposity

Table 2 showcases the associations of maternal fitness (VO_2peak_), gestational exercise volume, and pre-pregnancy BMI with aspects of MSC phenotype and infant adiposity. MSC respiration and lipid content were significantly and positively correlated to maternal average weekly (MET*min/wk) and total pregnancy exercise volume (Total MET*min) (p≤0.05), but not maternal pre-intervention fitness (VO_2peak_) or BMI (p>0.05). ASM and infant 1-month adiposity displayed negative correlation with ME volume during pregnancy and positive correlation with maternal pre-pregnancy BMI (p≤0.05). There were no associations between any of the maternal characteristics and infant 6-month body fat percentage (p>0.05, data not shown).

**Table 2.** Associations between maternal parameters and infant MSC function and body fat.

| **Maternal** | **Infant** | **r value** | **p value** |
| --- | --- | --- | --- |
| *16-week VO<sub>2peak</sub> (ml/kg/min)* | Intact MSC basal respiration | 0.29 | 0.14 |
|  | Intact MSC maximal respiration | 0.18 | 0.35 |
|  | MSC Oil RedO | 0.29 | 0.13 |
|  | MSC Acid soluble metabolites | -0.26 | 0.18 |
|  | 1-month body fat (%) | -0.40 | 0.06 |
| Average pregnancy Met*min/wk | Intact MSC basal respiration | 0.38 | **0.02** |
|  | Intact MSC maximal respiration | 0.43 | **0.01** |
|  | MSC Oil RedO | 0.47 | **<0.01** |
|  | MSC Acid soluble metabolites | -0.47 | **<0.01** |
|  | 1-month body fat (%) | -0.52 | **<0.01** |
| *Total pregnancy Met\*min* | Intact MSC basal respiration | 0.4 | **0.02** |
|  | Intact MSC maximal respiration | 0.43 | **0.01** |
|  | MSC Oil RedO | 0.48 | **0.05** |
|  | MSC Acid soluble metabolites | -0.41 | **0.01** |
|  | 1-month body fat (%) | -0.49 | **0.01** |
| Pre-pregnancy BMI | Intact MSC basal respiration | -0.14 | 0.37 |
|  | Intact MSC maximal respiration | -0.03 | 0.86 |
|  | MSC Oil RedO | -0.20 | 0.20 |
|  | MSC Acid soluble metabolites | 0.53 | **<0.001** |
|  | 1-month body fat (%) | 0.32 | 0.08 |

## Discussion

This study aimed to elucidate how ME affects offspring MSC insulin action, mitochondrial capacity and content, and fat oxidation and storage. Additionally, we did a 1- and 6-month follow up to determine if MSC metabolic changes are associated with lower infant adiposity. Here, we show that infant MSCs exposed to ME 1) have higher intact cell respiration, 2) are more insulin sensitive, 3) store more neutral lipids with higher fat oxidation, and 4) this MSC phenotype is associated with infant being leaner at 1-month, and adiposity at 6-months of age. Together, these data indicate that MSCs from exercising mothers display ‘classic’ adaptations typically seen with exercise training which contribute to reduce incidence of metabolic disease ^33^.

Son et al. ^34–36^ reported that mice from exercising mothers have higher whole body oxygen consumption rates at rest and during an acute bout of endurance exercise. These improvements were paralleled with greater mitochondrial content, PGC-1α expression, and AMPK phosphorylation in offspring skeletal muscle and brown adipose tissue^34^. In line with these findings, we found significantly higher intact cell basal and maximal respiration. Similarly, we found a significant increase in PGC-1α in the Ex-MSC group. This increase was seemingly driven by higher AMPK activation and ∼30% higher SIRT1 protein expression, considering the interdependence of SIRT1 and AMPK in transcriptional regulation of PGC-1α ^37^. Interestingly, despite the higher intact cell respiration and PGC-1α expression, we did not observe differences in the mitochondrial OXPHOS protein content, nor citrate synthase activity, pointing towards similar mitochondrial content between groups; however, it is worth noting that neither OXPHOS content nor CS-activity have been validated to correspond to mitochondrial enrichment within this cell model. Accordingly, it is hard to speculate if greater mitochondrial capacity is driven by greater mitochondrial content or respiratory capacity per mitochondrion.

To better understand if the higher cellular respiration is driven by the OXPHOS specific adaptations, we completed a series of permeabilized cell assays. To our surprise, we did not find any bioenergetic or complex activity differences between groups. Similar findings regarding the OXPHOS specific adaptations have been previously reported in pups of exercising dams ^7,38^. Specifically, ME before and during gestation did not alter mitochondrial complex I, complex II, or multi-substrate supported maximal OXPHOS or electron transport chain capacity in permeabilized muscle fibers. Considering that difference in both basal and maximal MSC respiration between groups was diminished after cell permeabilization, this would suggest a difference in regulatory control at the level of cellular cytosol, membrane transport or substrate availability; however, this is not likely considering that the increase in intact cell respiration upon FCCP addition was similar across groups (Figure 3B). It is more likely the difference in basal cellular respiration is due either to 1) a difference in substrate availability and/or preference, and/or 2) an intrinsically higher rate of energy demand in Ex-MSCs. In line with these hypotheses, 1) we observed higher lipid storage and oxidation (Figure 5), which could be a part of the observed difference in intact cell respiration. Stoichiometry of fatty acid oxidation favors electron input directly to the coenzyme Q pool, bypassing complex I which results in lower proton motive force generation and subsequently greater oxygen consumption for a given ΔG_ATP_. 2) State of a greater energetic demand (higher cellular AMP/ATP ratio) is reflected by higher AMPK activation in Ex-MSCs. Additionally, higher rate of ATP turnover would subsequently drive SIRT1 and PGC-1α adaptive response to increase mitochondrial flux and capacity in MSC from exercisers. While these are plausible mechanisms to explain greater basal intact cell respiration, further investigation is needed.

It is worth noting that despite any changes in the OXPHOS capacity, Quiclet et al. ^7^ reported greater mitochondrial affinity for fatty acid oxidation in rat offspring skeletal muscle exposed to exercise in utero. Specifically, ME led to an increase in palmitoyl-CoA, but not palmitoyl-carnitine affinity, suggestive of CPT-1 specific improvements. In our study, higher AMPK activation could translate to higher fat oxidation in Ex-MSCs by greater acetyl-CoA carboxylase phosphorylation, and subsequently lower CPT-1 inhibition. However, if this was the case, it would be reflected in the differential mitochondrial lipid availability between groups which was not observed. More likely is that association of AMPK activation and fatty acid oxidation to CO_2_ is reflective of greater energetic demand-driven fatty acid flux through TCA cycle. Herein, fatty acid influx and rate of β-oxidation is matched with higher flux of fatty acid-derived acetyl-CoAs through the TCA cycle in Ex-MSCs. Accordingly, this is reflected by lower accumulation of partially oxidized lipids (ASM production) and higher complete fatty acid oxidation to CO_2_ in Ex-MSCs, compared to Ctrl-MSCs. This would be on contrary to what has been reported in animal models and MSCs from mothers with obesity, where downregulation of lipid metabolism was associated with dysregulation of energy sensing pathways and AMPK activity ^2,4,39,40^. Improvements in offspring insulin action have been corroborated in animal models where ME improves offspring whole body glucose tolerance and insulin signaling ^20^. Similar to our findings in Ex-MSCs, two studies ^7,38^ found ME increases Akt activation and glucose uptake in pup skeletal muscle. It is worth noting that improvements in offspring insulin action were in parallel with increases in cell lipid storage. This is in line with the athlete’s ‘paradox’ theory ^41^, and more importantly showcases that greater neutral lipid storage is a factor of systems substrate preference and ability, rather than inability, to oxidize fatty acids.

ME was associated with improvements in infant body composition at 1-month of age which is in parallel with previous observations in animal models of ME ^7,34,42,43^. While we did not have a sufficient number of participants for a group comparison at the 6-month follow-up, we did observe significant correlations between infant MSC mitochondrial phenotype and adiposity. While a causal relationship between these parameters has not been established, it is important to note that MSC metabolic functioning precedes and therefore could influence infant growth trajectory. Accordingly, while it is difficult to speculate to what extent higher rate of energy turnover in MSCs determines infant adiposity, it is worth noting that multiple animal studies report lower adiposity gain in pups from exercise dams. Specifically, pups from exercisers had a significantly lower rate of fat and weight gain after a high fat diet ^7,42^, and this was in parallel with greater whole body respiration.

## Limitations

Current findings support and extend the growing evidence for promoting exercise during pregnancy. Strengths of the current study are a prospective, randomized controlled trial study design which provides the strongest evidence for causality. The use of fetal MSCs at birth eliminates some confounding variables (i.e., infant diet, environmental exposures, etc.), which may influence outcomes when using other methods. Further, we acknowledge that our sample consisted of ‘apparently healthy’ pregnant women reducing the generalizability of our findings; however, we had a diverse range of maternal BMIs to increase generalizability. Further, we did not account for participant dietary habits. Finally, we were not powered to control for maternal VO_2_ peak differences, and we did not obtain VO_2_peak from all the participants due to COVID-19 restrictions. This permits delineation between the effects of maternal pre-pregnancy exercise/fitness and maternal gestational exercise on infant MSC outcomes and body composition. Accordingly, this data is limited to interpretation that maternal gestational exercise and higher pre-pregnancy physical fitness are associated with improvements in MSC mitochondrial functioning and infant body composition. Future directions should aim to address these limitations and focus on delineating the effects of maternal exercise prior to and during pregnancy on infant metabolic health.

## Conclusion

ME induces an array of beneficial metabolic adaptations in offspring MSCs. Improvements in infant MSC insulin action and mitochondrial functioning seemingly influence infant body composition and could be protective of subsequent adiposity gain in early infancy. Collectively, by eliciting a multitude of positive metabolic adaptations in infant MSCs, ME could influence the trajectory of offspring metabolic health and reduce the transgenerational propagation of metabolic disease.

## Data Availability

Data generated/analyzed during the current study is available upon request from the corresponding or lead author.

## Clinical Trial

ClinicalTrials.gov Identifier: NCT03838146

