## Supplementary Table 1 for "Effects of maternal exercise on infant mesenchymal stem cell mitochondrial function, insulin action, and body composition in early infancy"

| **Supplementary Table 1. Western blot antibody information.** | | |
| --- | --- | --- |
| **Antibody** | **Manufacturer** | **Cat. No.** |
| Akt (Ser473) | Cell Signaling | 9271 |
| Akt (Thr308) | Cell Signaling | 4056 |
| Akt protein | Cell Signaling | 9272 |
| AMPK (Thr172) | Cell Signaling | 2531 |
| AMPK protein | Cell Signaling | 2532 |
| PGC1α | Abcam | ab106814 |
| SIRT1 | Cell Signaling | 2493 |
| Citrate Synthase | Abcam | ab96600 |
| Pyruvate Dehydrogenase | Cell Signaling | 2784 |
| Total OXPHOS | Abcam | ab9110411 |
| β-Actin | Cell Signaling | 4967 and 3700 |
