## Supplementary Table 2 for "Effects of maternal exercise on infant mesenchymal stem cell mitochondrial function, insulin action, and body composition in early infancy"

| **Supplementary Table 2. Maternal and Infant Characteristics** | | | | |
| --- | --- | --- | --- | --- |
| **Maternal Characteristics** | **AE (10)** | **CE (9)** | **RE (11)** | **p-value** |
| Age (yrs) | 30.5±4.6 | 28.7±1.8 | 31.6±3.0 | 0.18 |
| VO2peak (ml/kg/min)* | 25.2±3.2 | 24.9±6.0 | 22.5±5.9 | 0.54 |
| Average MET*min/wk between 16-36 weeks of gestation | 551.4±96.8 | 597.8±83.31 | 572±110.7 | 0.60 |
| Total MET*min between 16-36 weeks of gestation | 11785±4413 | 13913±2932 | 12572±2473 | 0.4 |
| Pre-pregnancy BMI | 23.6±2.3 | 25.6±4.5 | 24.7±4.5 | 0.52 |
| 16-week BMI | 24.8±2.7 | 26.7±4.8 | 26.1±5.1 | 0.64 |
| 16-week Body Fat (%) | 35.1±3.2 | 33.5±4.5 | 33.2±4.6 | 0.58 |
| 16-week Total Cholesterol (mg/dl) | 184.6±27.1 | 159.4±20.2 | 184.9±37.6 | 0.13 |
| 16-week LDL (mg/dl) | 108.9±31.2 | 78.4±16.7 | 96.6±31.4 | 0.08 |
| 16-week HDL (mg/dl) | 52.3±19.1 | 62.7±7.9 | 69.1±15.4 | **0.05** |
| 16-week non-HDL (mg/dl) | 132.3±30.9 | 97.1±18.5 | 115.3±34.6 | 0.06 |
| 16-week Triglycerides (mg/dl) | 116.6±46.3 | 93.2±31.9 | 92.2±29.3 | 0.27 |
| 16-week Glucose (mg/dl) | 78.4±6.9 | 79.1±6.9 | 78.4±6.2 | 0.96 |
| 16-week Lactate (mmol/l) | 1.1±0.4 | 0.9±0.3 | 0.9±0.6 | 0.73 |
| OGTT (mg/dL) | 96.5±23.3 | 113.1±34.2 | 116.6±20.4 | 0.24 |
| Gestational weight gain (kg) | 15.4±4.3 | 22.7±19.9 | 21.5±14.8 | 0.48 |
| Gestational length (weeks) | 39.8±1.1 | 39.7±1.1 | 39.2±1.4 | 0.53 |
| Parity | 1 (0, 2) | 0 (0, 1) | 1 (0, 3) | 0.66 |
| Mode of delivery (SVD/C-section) | 2 (1, 2) | 2 (1, 2) | 2 (1, 2) | 0.93 |
| **Infant Characteristics** | **AE (10)** | **CE (9)** | **RE (11)** | **p-value** |
| Fetal sex (F/M) | 3/7 | 3/6 | 3/8 | 0.96 |
| Birth weight (kg) | 3.6±.4 | 3.5±.5 | 3.4±.4 | 0.56 |
| Birth length (m) | 0.49±0.03 | 0.49±0.01 | 0.50±0.03 | 0.59 |
| Birth BMI | 14.7±1.6 | 14.7±2.0 | 13.5±1.2 | 0.13 |
| Head circumference (m) | 0.34±0.01 | 0.35±0.02 | 0.34±0.01 | 0.52 |
| Chest circumference (m) | 0.33±0.02 | 0.33±0.02 | 0.33±0.01 | 0.9 |
| Abdominal circumference (m) | 0.31±0.02 | 0.31±0.02 | 0.31±0.01 | 0.98 |
| Apgar-1 minute | 8 (7, 9) | 9 (8, 9) | 8 (7, 9) | 0.79 |
| Apgar-5 minute | 9 (9, 9) | 9 (9, 9) | 9 (9, 9) | 0.99 |
| All data expressed as mean ± SD, t-test, p≤0.05. * VO2peak n=22/30 due to COVID; OGTT = 1-hour blood oral glucose tolerance test at 24-28 weeks; Gestational weight gain = delivery - pre-pregnancy; SVD = spontaneous vaginal delivery; F= female infant sex; M = male infant sex. | | | | |
