## Supplementary Figure 1 for "Effects of maternal exercise on infant mesenchymal stem cell mitochondrial function, insulin action, and body composition in early infancy"

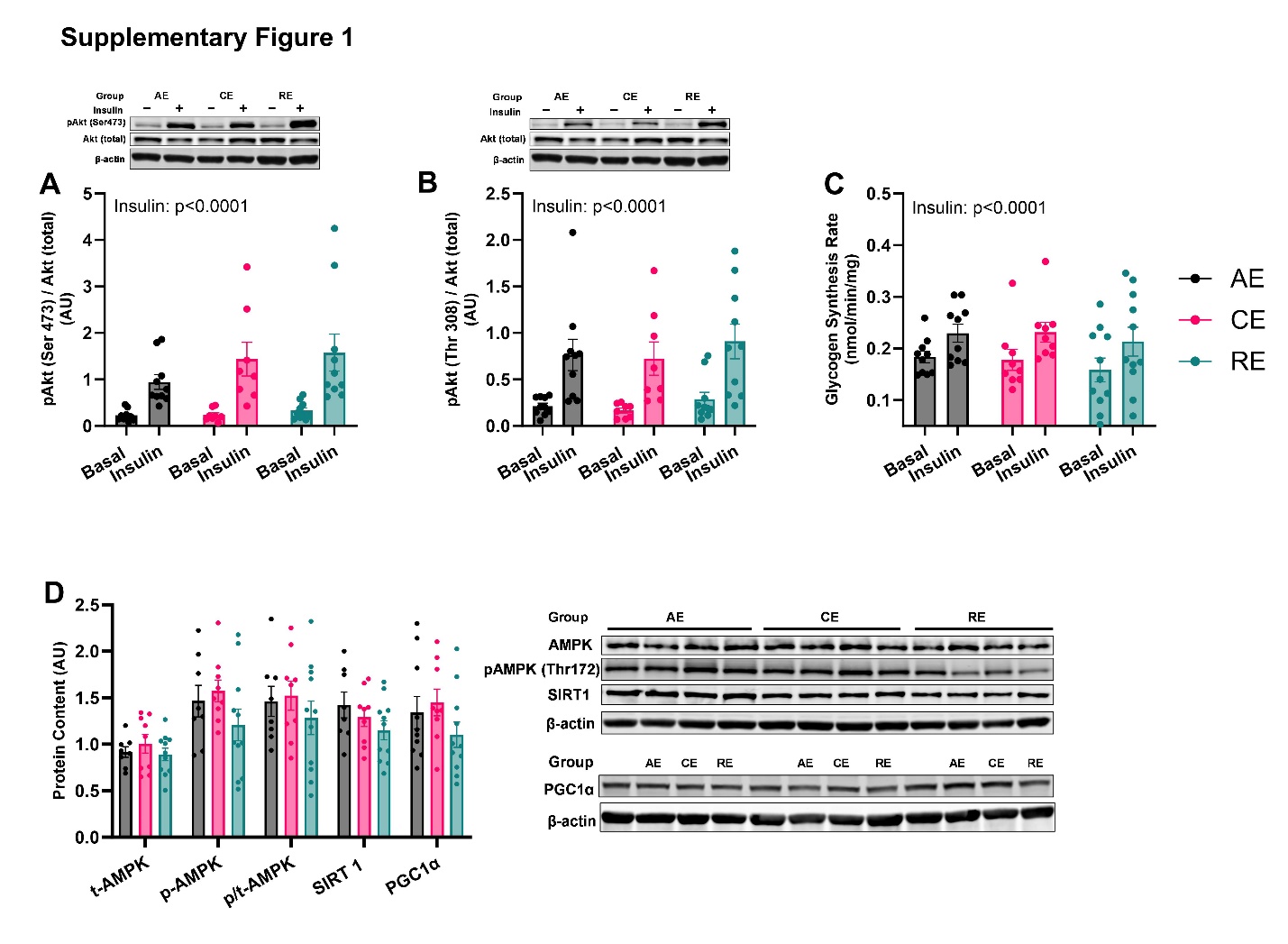
**Supplementary Figure 1. Insulin action and expression of redox-sensitive and energy-sensing proteins.** Insulin-stimulated Akt phosphorylation at Ser473 (A) and Thr308 (B) was similar between exercise groups. Both absolute rates and relative increases in glycogen synthesis in response to insulin were similar across groups (C). Expression and activation of AMPK, SIRT1, and PGC1-α (D). Data expressed as mean ± SEM. n=8-10/group. Two-way repeated measures ANCOVA (covariate – HDL at 16 weeks) and One-way ANCOVA. AE- aerobic; CE – combination; RE – resistance group.
