## Supplementary Figure 2 for "Effects of maternal exercise on infant mesenchymal stem cell mitochondrial function, insulin action, and body composition in early infancy"

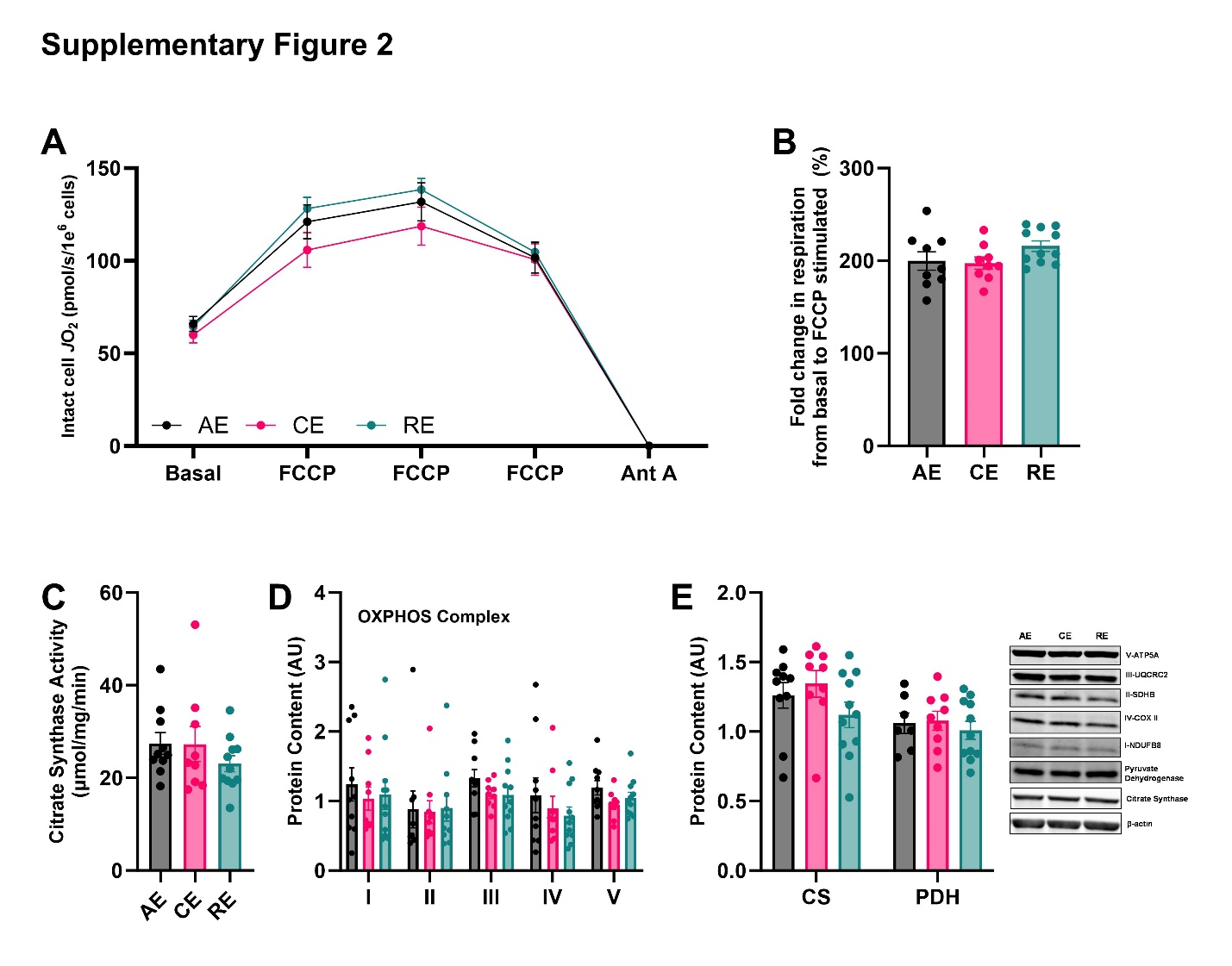
**Supplementary Figure 2. Intact cell respiration and mitochondrial content.** Intact cell respiration (A) and response to FCCP (B) were similar between groups. Citrate synthase activity (C) was not different between groups. Protein expression of OXPHOS (D), citrate synthase (CS), and pyruvate dehydrogenase (PDH) (E) was similar between exercise groups. Data expressed as mean ± SEM. n=8-10/group. ANCOVA (covariate – HDL at 16 weeks). AE- aerobic; CE – combination; RE – resistance group.
