## Supplementary Figure 3 for "Effects of maternal exercise on infant mesenchymal stem cell mitochondrial function, insulin action, and body composition in early infancy"

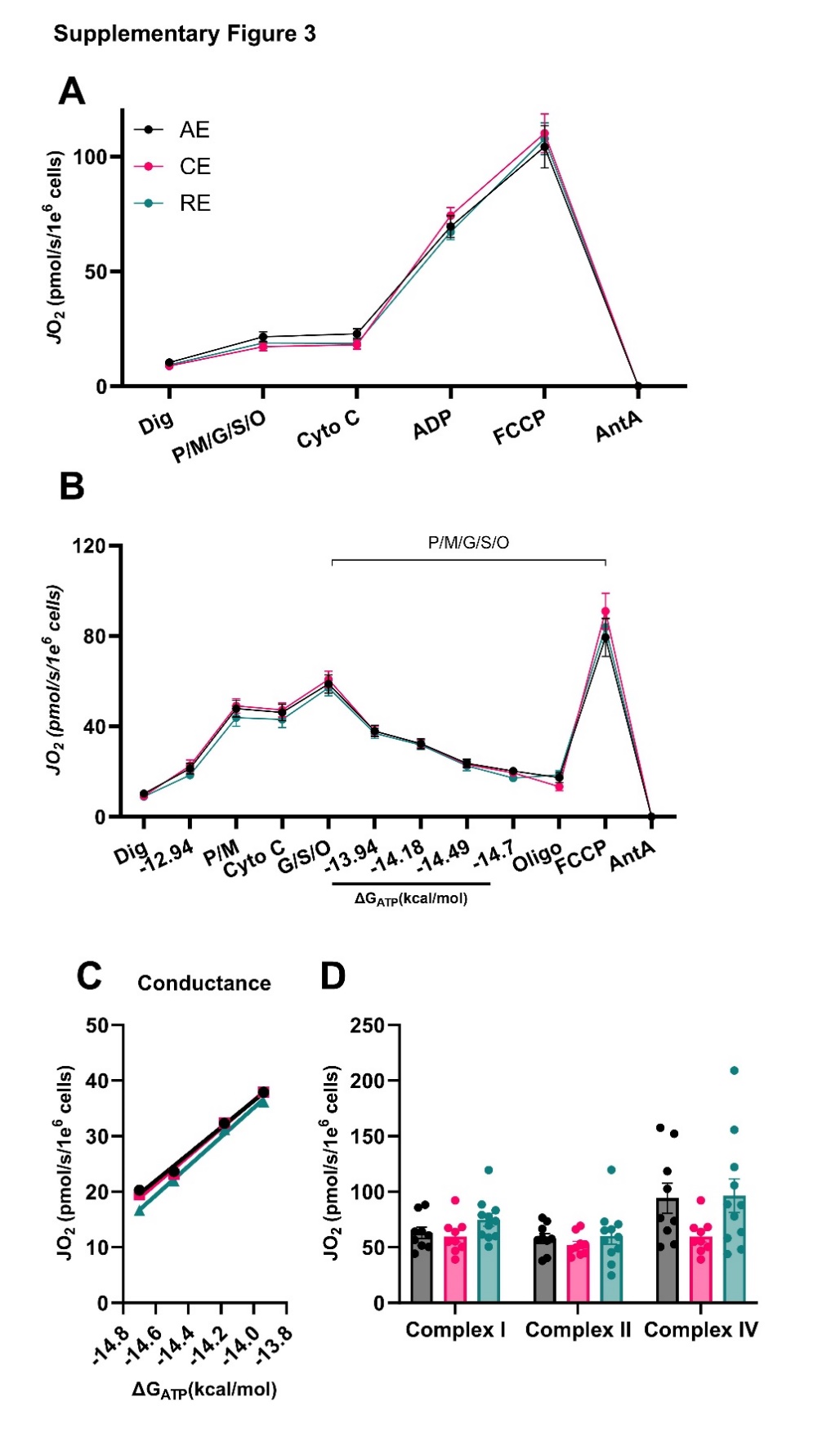
**Supplementary Figure 3. Permeabilized cell respiration.** ADP and FCCP-stimulated respiration in permeabilized cells were similar between exercise groups (A), regardless of the presence of ΔGATP (B). Conductance (C). Cellular respiration supported with complex I, II, or IV specific substrates (D). Data expressed as mean ± SEM. n=8-10/group. ANCOVA (covariate – HDL at 16-weeks). AE- aerobic; CE – combination; RE – resistance group.
