## Supplementary Figure 4 for "Effects of maternal exercise on infant mesenchymal stem cell mitochondrial function, insulin action, and body composition in early infancy"

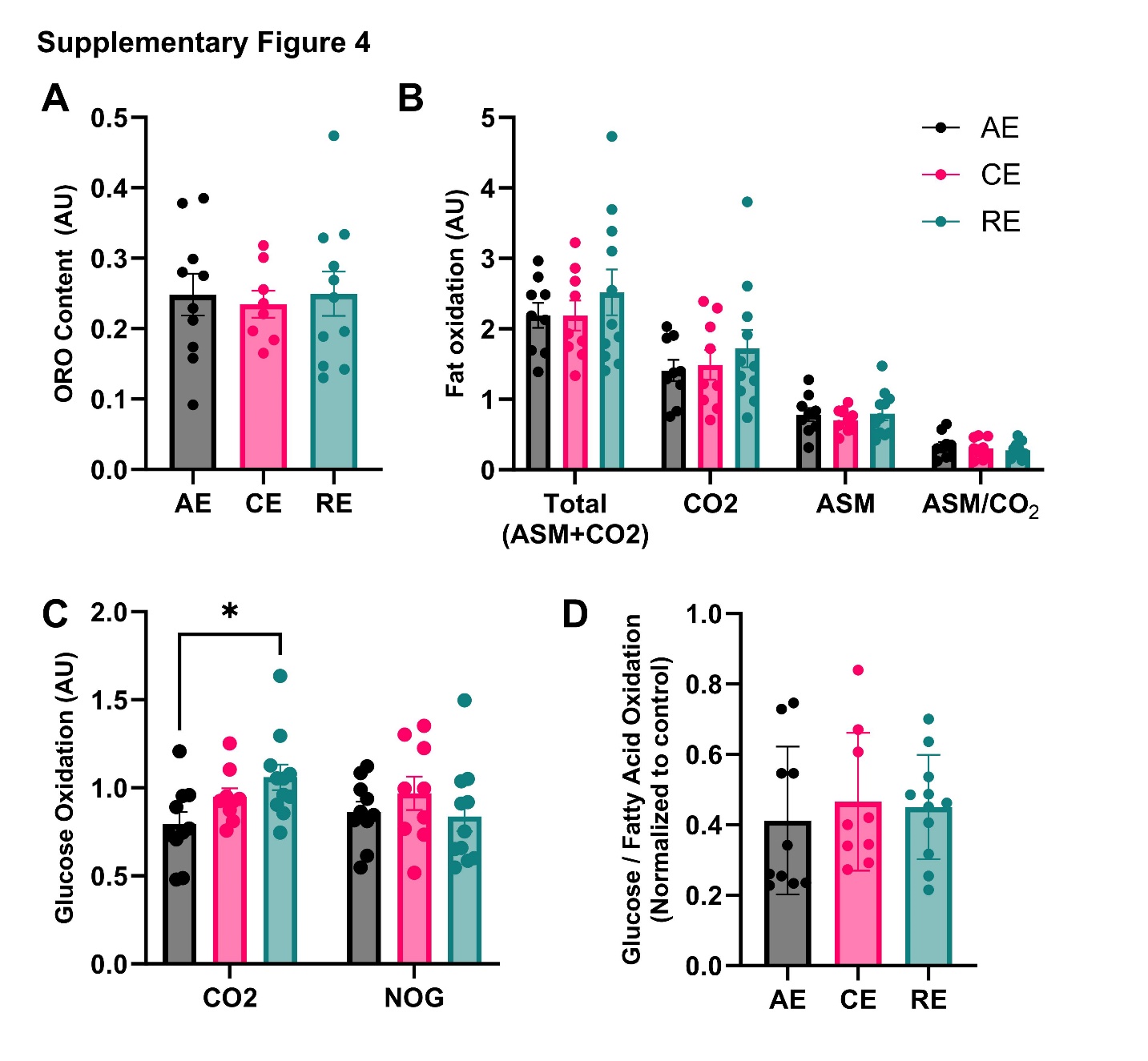

**Supplementary Figure 4. Fat and glucose oxidation, and fat storage.** Neutral lipid storage measured by oil-red-o stain was similar between exercise groups (A). Fatty acid uptake (total), complete oxidation (CO_2_), incomplete oxidation (ASM), and partitioning ratio (ASM/CO_2_) were similar between exercise groups (B). Glucose complete oxidation was higher in the resistance group compared to the aerobic group, without any difference in the rate of non-oxidized glycolysis (NOG) between groups (C). Despite higher complete oxidation in RE compared to AE, ratio of glucose/fatty acid oxidation was similar across groups (D). Data expressed as mean ± SEM. n=8-10/group. ANCOVA (covariate – HDL at 16 weeks), with Bonferroni post-hoc test. AE- aerobic; CE – combination; RE – resistance group.
