## Supplementary Figure 5 for "Effects of maternal exercise on infant mesenchymal stem cell mitochondrial function, insulin action, and body composition in early infancy"

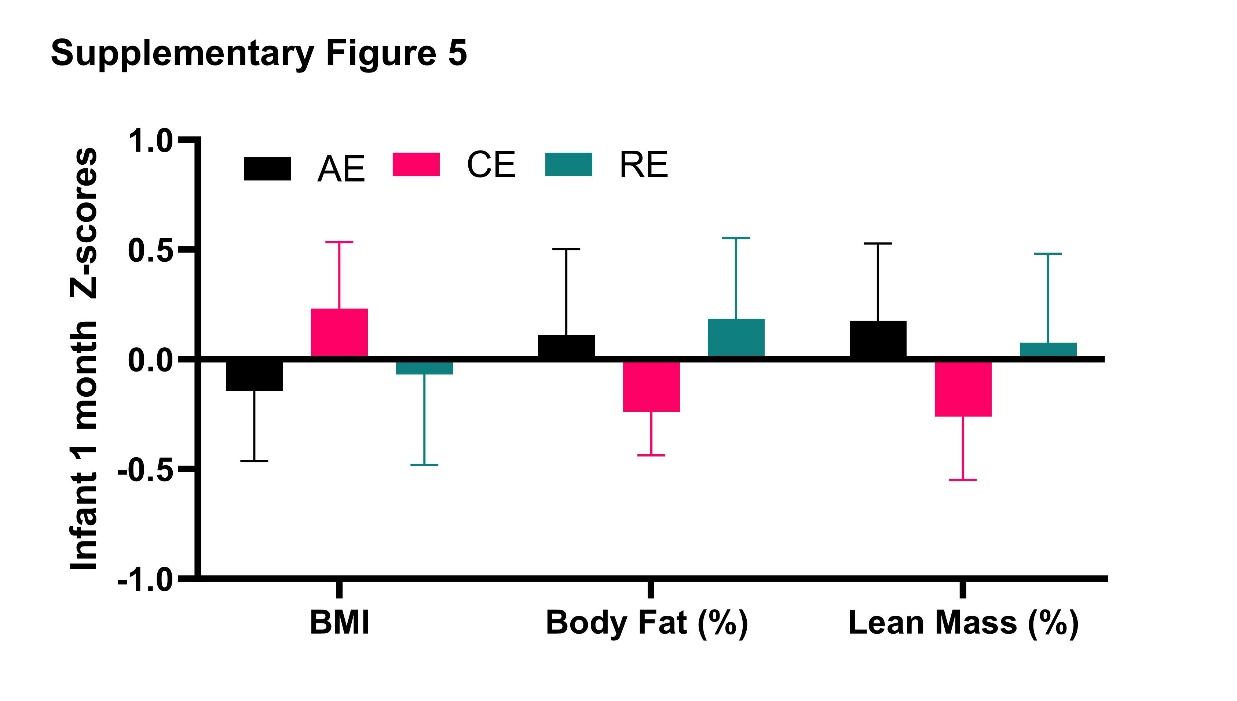
**Supplementary Figure 5. Infant body composition.** BMI, body fat percentage, and lean mass z scores were similar across exercise groups. Data expressed as mean ± SEM. n=8-10/group. ANCOVA (covariate – HDL at 16 weeks). AE- aerobic; CE – combination; RE – resistance group.
