## Supplementary Figure 6 for "Effects of maternal exercise on infant mesenchymal stem cell mitochondrial function, insulin action, and body composition in early infancy"

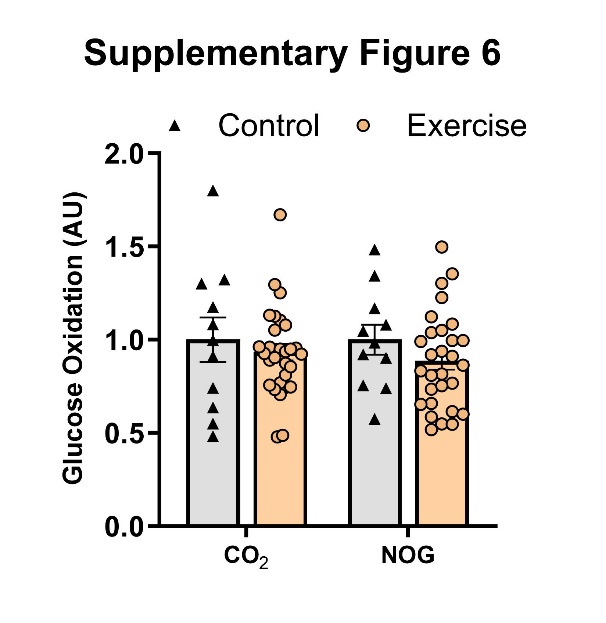
**Supplementary Figure 6.** Glucose oxidation to CO_2_ and the rate of non-oxidized glycolysis were similar between the control and combined exercise group (B). Data expressed as mean ± SEM. control n=11, exercise, n=30; * p≤0.05.
